# Anti-nuclear matrix protein 2 antibody-positive idiopathic inflammatory myopathies represent extensive myositis without dermatomyositis-specific rash

**DOI:** 10.1101/2021.02.10.21251468

**Authors:** Yuki Ichimura, Miwako Shobo, Sae Inoue, Mari Okune, Akemi Maeda, Ryota Tanaka, Noriko Kubota, Isao Matsumoto, Akiko Ishii, Akira Tamaoka, Asami Shimbo, Masaaki Mori, Tomohiro Morio, Takayuki Kishi, Takako Miyamae, Jantima Tanboon, Michio Inoue, Ichizo Nishino, Manabu Fujimoto, Toshifumi Nomura, Naoko Okiyama

## Abstract

**Objective:** Myositis-specific autoantibodies (MSAs) define distinct clinical subsets of idiopathic inflammatory myopathies (IIMs). The anti-nuclear matrix protein 2 (NXP2) antibody, a MSA detected in juvenile/adult IIMs, has been reported to be associated with a high risk of subcutaneous calcinosis, subcutaneous edema, and internal malignancies. The study aimed to clarify the clinical features of anti-NXP2 antibody-positive IIMs in detail.

**Methods:** This multi-center retrospective observational study on 76 anti-NXP2 antibody-positive patients. The antibody was detected via a serological assay using immunoprecipitation and western blotting. The patients were selected from 162 consecutive Japanese patients with IIMs.

**Results:** The cohort of anti-NXP2 antibody-positive IIMs included 29 juvenile patients and 47 adult patients. Twenty-seven (35.5%) patients presented with polymyositis phenotype without dermatomyositis-specific skin manifestations (heliotrope rash and Gottron sign/papules); this was more common in the adults than children (48.9% vs. 15.8%, *P* < 0.01). Nine (11.8%) patients had subcutaneous calcinosis, and 20 (26.3%) patients had subcutaneous edema. In addition, the proportion of patients with muscle weakness extending to the distal limbs was high (36 patients [47.4%]) in this cohort. Adult patients had a higher prevalence of malignancy than the general population (age-standardized incidence ratio of malignancies: 22.4).

**Conclusion:** Anti-NXP2 antibody-positive IIMs, which include dermatomyositis sine dermatitis, are characterized by atypical skin manifestations and extensive muscular involvement.

## Introduction

Inflammatory idiopathic myopathies (IIMs) are mainly characterized by dermatomyositis (DM) and polymyositis (PM). Myositis is characterized by a predominance in the proximal limb and trunk muscles with/without DM-specific skin manifestations such as heliotrope rash and Gottron sign/papules (1). Myositis-specific antibodies (MSAs) have been identified recently in the sera of patients with DM/PM (2), and different MSAs are associated with distinct clinical subsets. Anti-nuclear matrix protein 2 (NXP2) antibody belongs to MSAs, and was first detected in the sera of patients with juvenile DM as an anti-MJ antibody (3). Notably, the population with anti-NXP2 antibody is the second largest (20%) among all juvenile DM cases (4). Moreover, 5%–30% of adult DM cases are positive for anti-NXP2 Antibody (5, 6).

Some clinical characteristics of anti-NXP2 antibody-positive IIMs have been suggested. Although subcutaneous calcinosis and edema have been observed frequently in cases of anti-NXP2 antibody-positive DM (5), anti-NXP2 antibody was also detected in the sera of patients with DM sine dermatitis (DMSD) in whom skin manifestations were not observed, but the muscle pathology suggested DM (7). A high risk of malignancy has also been suggested, especially in anti-NXP2 antibody-positive adults (5,6). However, the characteristics of patients with anti-NXP2 antibody-positive IIMs clinically diagnosed as PM have not been evaluated in detail, and the differences between juvenile and adult patients with anti-NXP2 antibody-positive IIMs were scarcely reported. Therefore, this study aimed to clarify the clinical features of IIMs, including DM and PM, in anti-NXP2 antibody-positive patients.

## Patients and methods

### Patients

One-hundred fifty-six consecutive Japanese patients with IIMs were identified. All the patients fulfilled the 2017 European League of Rheumatology (EULAR)/American College of Rheumatology (ACR) classification criteria for juvenile and adult IIMs (1) and had been treated at University of Tsukuba Hospital and our collaborating medical centers (Supplemental list) from January 2017 to December 2020. The sera of these patients were negative for anti-transcription intermediatory factor (TIF) 1γ, anti-melanoma differentiation-associated protein 5 (MDA5), anti-Mi-2, and anti-aminoacyl transfer RNA synthetase antibodies, tested using commercially available enzyme-linked immunosorbent assay kits (Medical & Biological Laboratories, Nagoya, Japan). There were 14 patients with anti-small ubiquitin-like modifier activation enzyme (SAE) 1/2 antibody and one patient with anti-TIF1β antibody that were excluded from the study. The findings in all patients were confirmed by immunoprecipitation and western blotting (IP-WB). A total of 76 anti-NXP2 antibody-positive patients were included in this study. The anti-NXP2 antibody was detected by IP-WB. No autoantibodies/MSAs were detected in the serum samples of the remaining 56 patients. Clinical information was collected retrospectively by reviewing patients’ medical charts. This study was approved by the ethics committee of University of Tsukuba Hospital (H29-111) in accordance with the Helsinki Declaration.

### IP-WB

Twenty microliters of serum from each patient were mixed with 4 mg of protein A-Sepharose beads (Cytiva, Marlborough, MA) in IPP buffer (10 mM Tris-HCl, pH 8.0, 50 mM NaCl, 0.1% 4-nonylphenyl-polyethylene glycol [BioVision, Milpitas, CA]) for 2 h. Antibody-bound sepharose beads were washed with IPP buffer and incubated with extracts of 1 × 10^7^ K562 cells (ATCC, Manassas, VA) at 4°C for 2 h. The precipitated proteins were fractionated by sodium dodecyl sulfate-polyacrylamide gel electrophoresis using 10% polyacrylamide gel and then transferred onto nitrocellulose membranes using the Mini Trans-Blot^®^ Cell (Bio-Rad, Hercules, CA). The membranes were blocked with 5% skim milk, incubated with murine anti-human NXP2 monoclonal antibody (clone 17A9, Medical & Biological Laboratories), mouse anti-human SAE1 monoclonal antibody (clone 1G4–1G5, Abnoba, Taipei City, Taiwan), and anti-human SAE2 polyclonal antibody (BETHYL Laboratories, Montgomery, TX) or murine anti-human TIF1β monoclonal antibody (clone 1Tb3, Merck, Darmstadt, Germany) overnight at 4°C. They were then incubated with peroxidase-labeled goat anti-mouse IgG polyclonal antibodies (Santa Cruz Biotechnology, Dallas, TX) for NXP2, SAE1, and TIF1β, or goat anti-rabbit IgG polyclonal antibodies (Santa Cruz Biotechnology) for SAE2 after washing with tris-buffered saline with Tween 20 (ThermoFisher Scientific, Franklin, MA).

### Statistical analysis

Statistical analyses were performed using SPSS version 22 (IBM, North Castle, NY). A *P*-value of <0.05 was considered statistically significant.

## Results

### Clinical characteristics of patients of anti-NXP2 Antibody-positive IIMs

The clinical characteristics of 76 patients with anti-NXP2 antibody are summarized in Table 1. The age distribution was bimodal, including 29 juvenile patients (38.2%; mean age at onset [range], 8 [2–14] years) and 47 adult patients (61.8%; 52 [18–82] years). Twenty-seven (35.5%) patients presented with PM without DM-specific skin manifestations (heliotrope rash and Gottron sign/papules). Of these 27 patients, eight patients (10.5%) did not have any rashes and 19 (25.0%) patients presented other non-specific skin manifestations (Figure 1A). Nine (11.8%) patients had subcutaneous calcinosis, and 20 (26.3%) patients had subcutaneous edema.

**Table 1.**
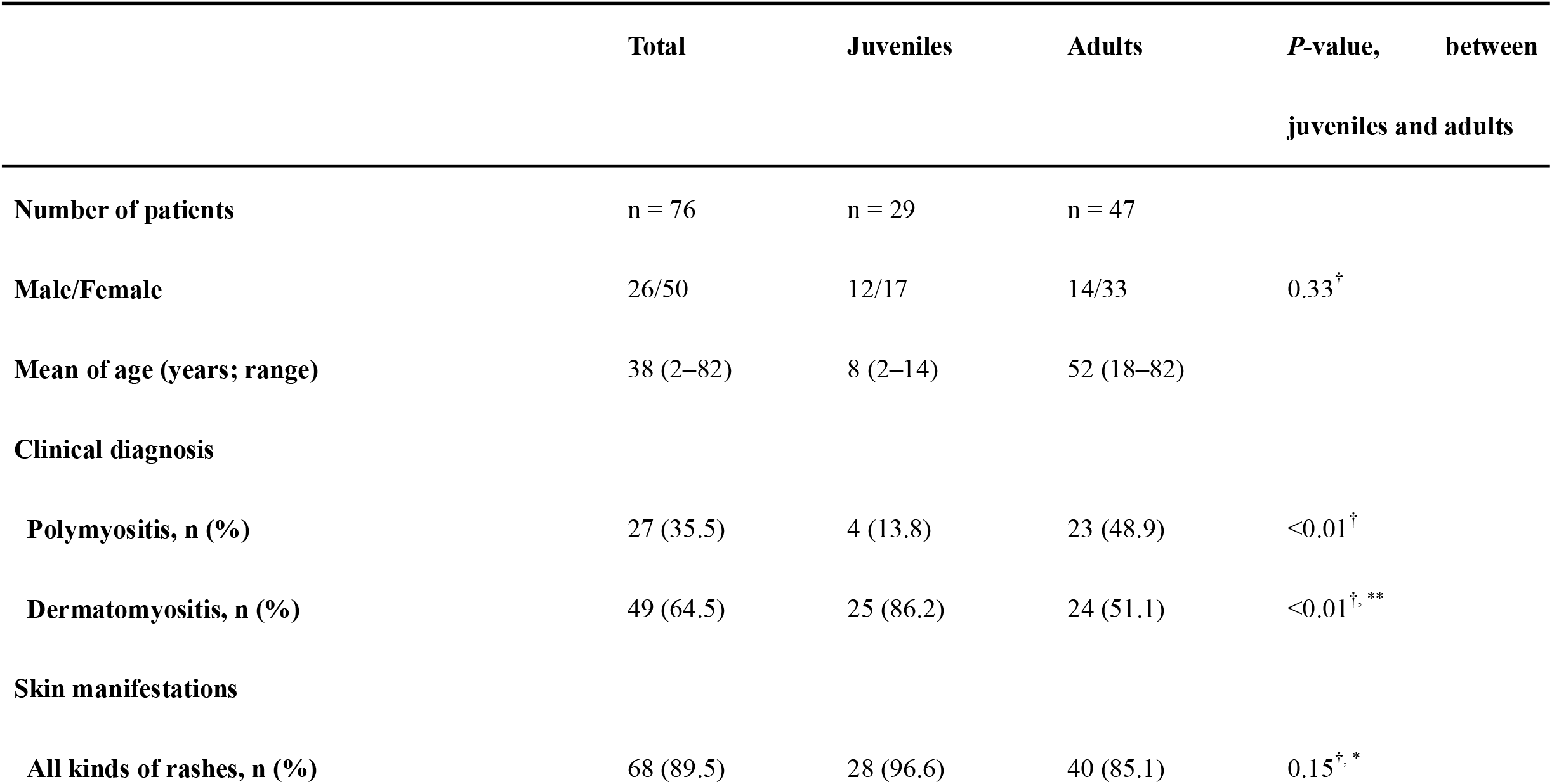
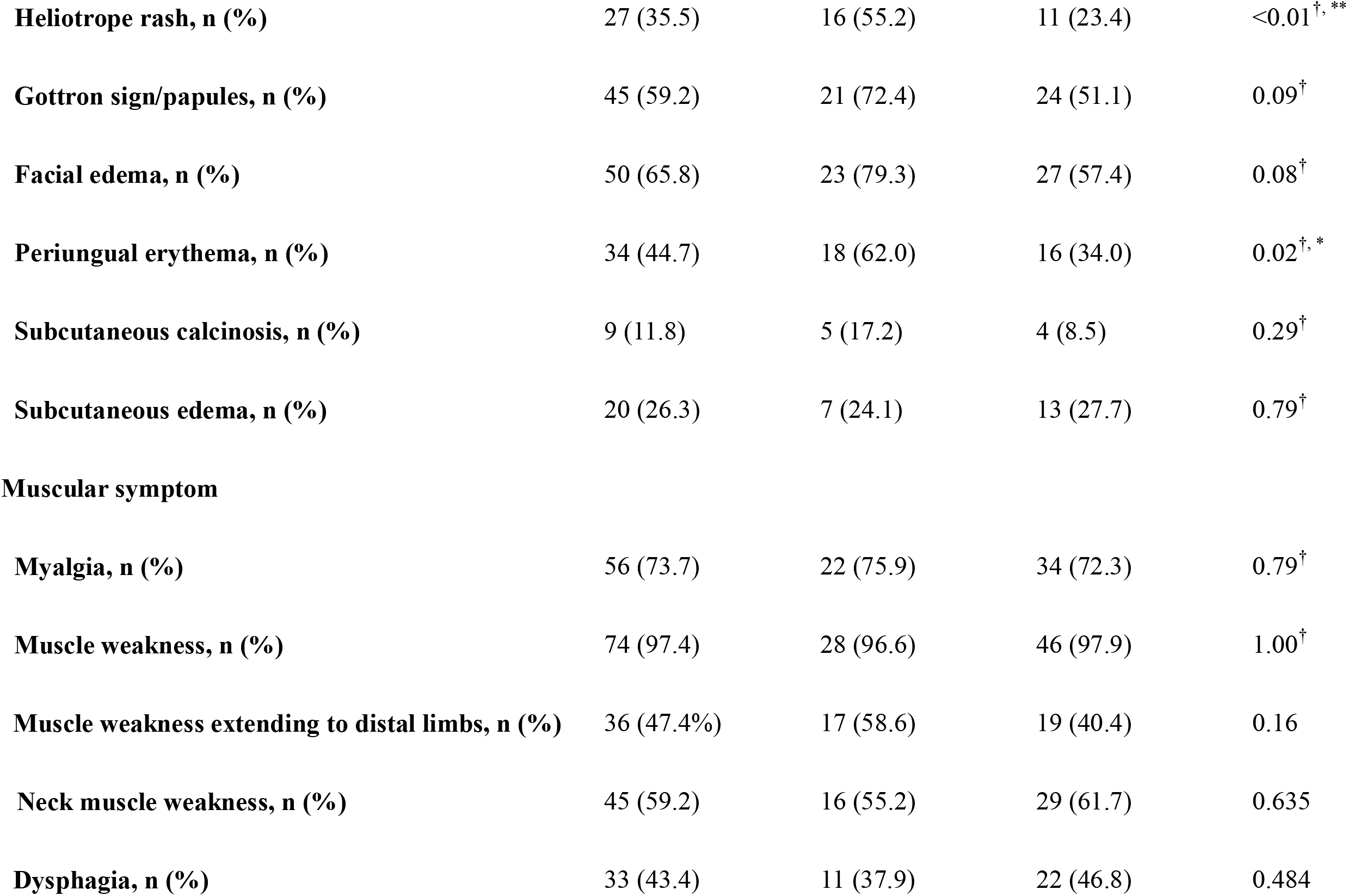
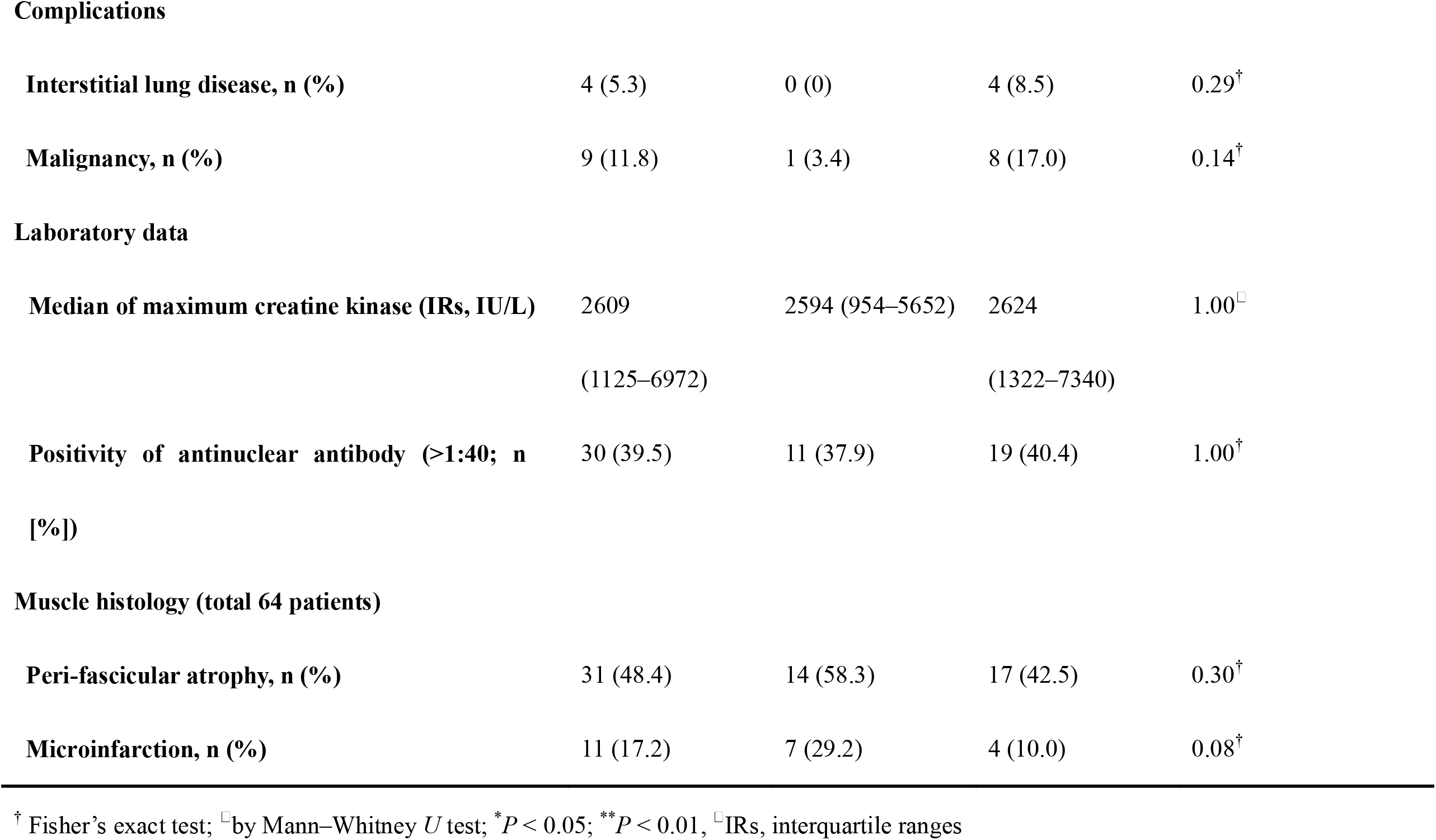
Clinical and laboratory characteristics of patients with anti-nuclear matrix protein 2 antibody-positive idiopathic inflammatory myopathies.

|  | **Total** | **Juveniles** | **Adults** | ***P*-value, between juveniles and adults** |
| --- | --- | --- | --- | --- |
| **Number of patients** | n = 76 | n = 29 | n = 47 |  |
| **Male/Female** | 26/50 | 12/17 | 14/33 | 0.33<sup>†</sup> |
| **Mean of age (years; range)** | 38 (2–82) | 8 (2–14) | 52 (18–82) |  |
| **Clinical diagnosis** |  |  |  |  |
| **Polymyositis, n (%)** | 27 (35.5) | 4 (13.8) | 23 (48.9) | <0.01<sup>†</sup> |
| **Dermatomyositis, n (%)** | 49 (64.5) | 25 (86.2) | 24 (51.1) | <0.01<sup>†, **</sup> |
| **Skin manifestations** |  |  |  |  |
| **All kinds of rashes, n (%)** | 68 (89.5) | 28 (96.6) | 40 (85.1) | 0.15<sup>†, *</sup> |
| **Heliotrope rash, n (%)** | 27 (35.5) | 16 (55.2) | 11 (23.4) | <0.01<sup>†, **</sup> |
| **Gottron sign/papules, n (%)** | 45 (59.2) | 21 (72.4) | 24 (51.1) | 0.09<sup>†</sup> |
| **Facial edema, n (%)** | 50 (65.8) | 23 (79.3) | 27 (57.4) | 0.08<sup>†</sup> |
| **Periungual erythema, n (%)** | 34 (44.7) | 18 (62.0) | 16 (34.0) | 0.02<sup>†, *</sup> |
| **Subcutaneous calcinosis, n (%)** | 9 (11.8) | 5 (17.2) | 4 (8.5) | 0.29<sup>†</sup> |
| **Subcutaneous edema, n (%)** | 20 (26.3) | 7 (24.1) | 13 (27.7) | 0.79<sup>†</sup> |
| **Muscular symptom** |  |  |  |  |
| **Myalgia, n (%)** | 56 (73.7) | 22 (75.9) | 34 (72.3) | 0.79<sup>†</sup> |
| **Muscle weakness, n (%)** | 74 (97.4) | 28 (96.6) | 46 (97.9) | 1.00<sup>†</sup> |
| **Muscle weakness extending to distal limbs, n (%)** | 36 (47.4%) | 17 (58.6) | 19 (40.4) | 0.16 |
| **Neck muscle weakness, n (%)** | 45 (59.2) | 16 (55.2) | 29 (61.7) | 0.635 |
| **Dysphagia, n (%)** | 33 (43.4) | 11 (37.9) | 22 (46.8) | 0.484 |
| Complications |  |  |  |  |
| **Interstitial lung disease, n (%)** | 4 (5.3) | 0 (0) | 4 (8.5) | 0.29<sup>†</sup> |
| **Malignancy, n (%)** | 9 (11.8) | 1 (3.4) | 8 (17.0) | 0.14<sup>†</sup> |
| Laboratory data |  |  |  |  |
| **Median of maximum creatine kinase (IRs, IU/L)** | 2609<br>(1125–6972) | 2594 (954–5652) | 2624<br>(1322–7340) | 1.00<sup>□</sup> |
| **Positivity of antinuclear antibody (>1:40; n [%])** | 30 (39.5) | 11 (37.9) | 19 (40.4) | 1.00<sup>†</sup> |
| Muscle histology (total 64 patients) |  |  |  |  |
| **Peri-fascicular atrophy, n (%)** | 31 (48.4) | 14 (58.3) | 17 (42.5) | 0.30<sup>†</sup> |
| **Microinfarction, n (%)** | 11 (17.2) | 7 (29.2) | 4 (10.0) | 0.08<sup>†</sup> |

<sup>†</sup> Fisher's exact test; <sup>□</sup> by Mann–Whitney *U* test; \**P* < 0.05; \*\**P* < 0.01, <sup>□</sup>IRs, interquartile ranges

**Table 2.**
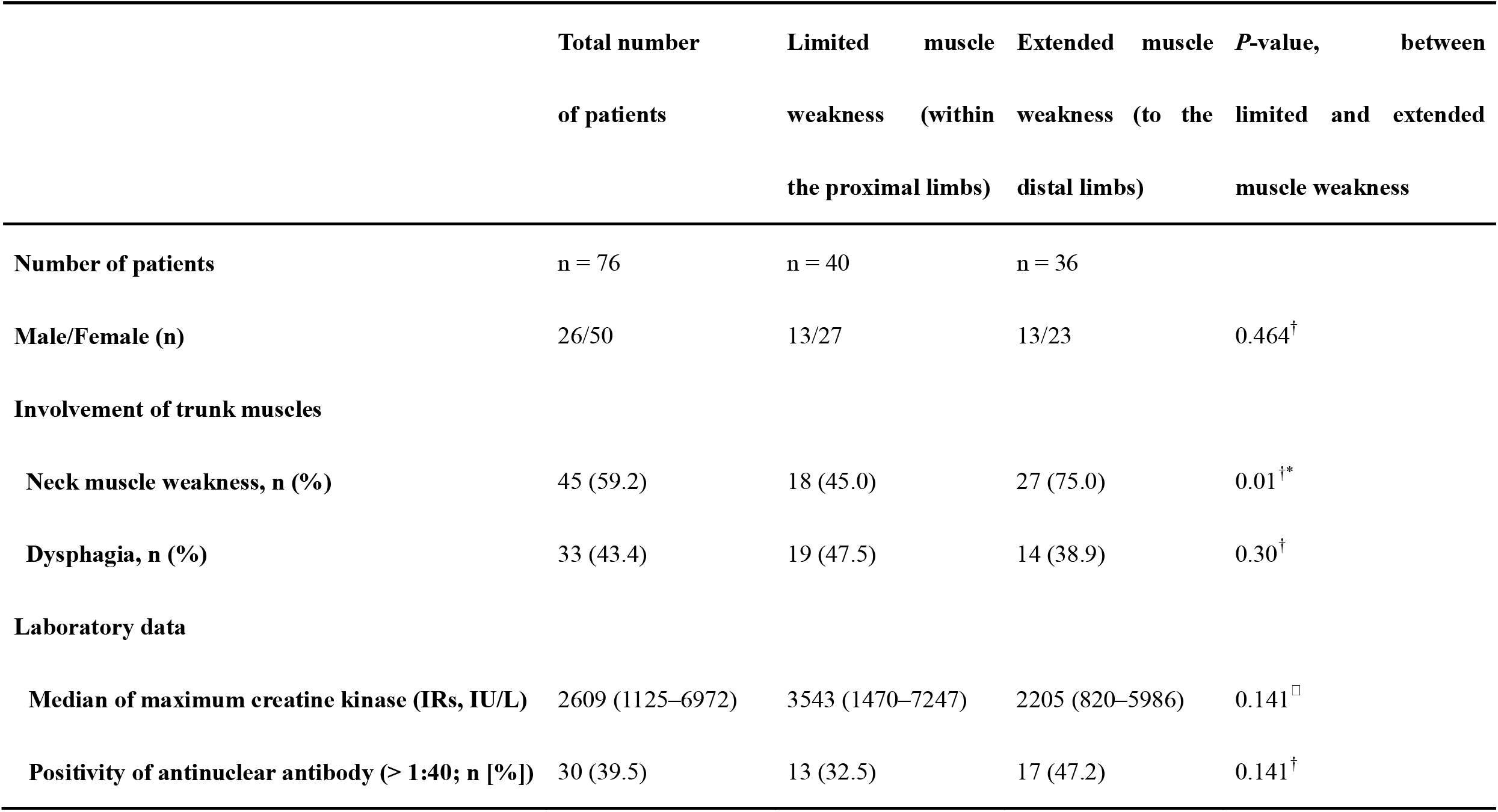
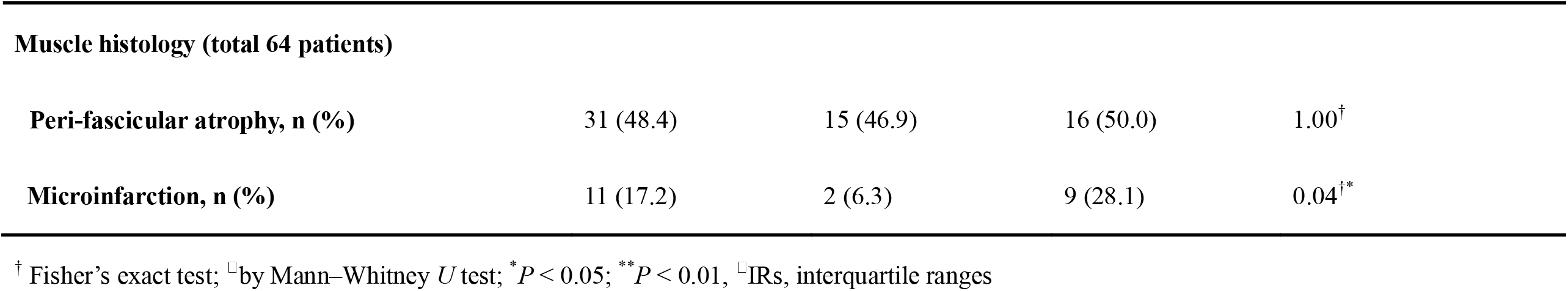
Muscular involvement in patients with anti-nuclear matrix protein 2 antibody-positive idiopathic inflammatory myopathies.

**Figure 1.**
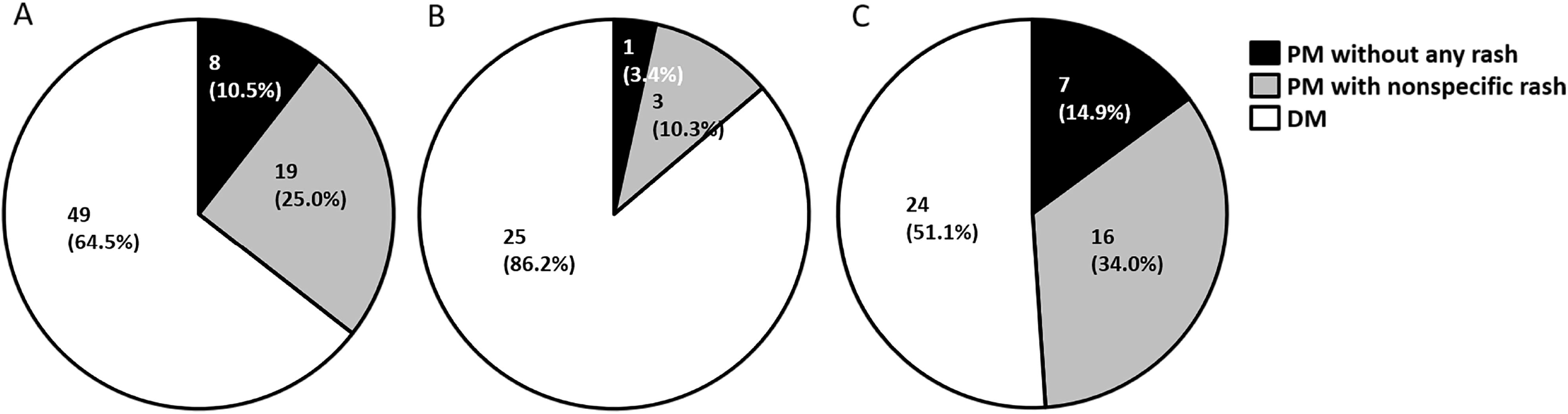
Polymyositis and dermatomyositis phenotypes in anti-nuclear matrix protein 2 antibody-positive idiopathic inflammatory myopathies. **(A)** Total cases, **(B)** Juvenile cases, and **(C)** adult cases. The number of cases (percentages) is presented in the graphs. PM, polymyositis; DM, dermatomyositis.

All patients except one had muscle pain and/or muscle weakness. One patient did not complain of any muscular symptoms but had an elevated serum creatine kinase (CK) level. Thirty-six (47.4%) patients demonstrated muscle weakness of the distal limbs in addition to that of the proximal limbs. Moreover, 45 (59.2%) patients showed neck muscle weakness, and 33 (43.4%) patients developed dysphagia. Muscle biopsy samples were histologically evaluated in 64 patients—31 (48.4%) patients had perifascicular atrophy (PFA) and 11 (17.2%) patients had evident microinfarction. The median (interquartile range [IR]) CK level was 2609 (1125–6972) IU/L, with six patients (three juveniles and three adults) having normal serum CK levels.

Four (5.3%) patients, including two with DM phenotype and two with PM phenotype, showed interstitial lung disease (ILD). Nine (11.8%) patients, including three with DM phenotype and six with PM phenotype, had malignancy.

Antinuclear antibody (ANA) was positive (titer >1:40) in 30 (39.5%) patients; the ANA titer was 1:80, 1:160, 1:320, 1:640, 1:1280, and >1:2560 in nine, four, eight, six, two, and one patients, respectively.

### Comparison between juvenile and adult patients

Adult patients with anti-NXP2 antibody-positive IIMs more frequently presented the PM phenotype than juvenile patients (n = 23, 48.9% vs. n = 4, 13.8%; *P* < 0.01 by Fisher’s exact test; Table 1; Figure 1B and 1C). In other words, DM-specific skin manifestations were more prevalent among the juvenile patients than among adult patients (n = 25, 86.2% vs. n = 24, 51.1%; *P* < 0.01; Table 1; Figure 1B and 1C). In particular, heliotrope rashes were more frequently observed in juvenile patients than in adults (n = 16, 55.2% vs. n = 11, 23.4%; *P* < 0.01 by Fisher’s exact test). Periungual erythema was also more common in juvenile patients than in adult patients (n = 18, 62.0% vs. n = 16, 34.0%; *P* = 0.02 by Fisher’s exact test).

Muscular manifestations were not significantly different between juvenile and adult patients. The median CK levels were also similar between juvenile and adult patients (2596 vs. 2624 IU/L; *P* = 1.00 by Mann–Whitney *U* test).

Four (8.5%) adult patients had ILD, while no juvenile patients had ILD (0%; *P* = 0.29 by Fisher’s exact test). Malignancies were diagnosed in eight (17.0%) adult patients, including cervical cancer in two patients, colon cancer in one patient, gastric cancer in two patients, breast cancer in one patient, uterine cancer in one patient, and cutaneous squamous cell carcinoma on the cheek in one patient. Because the age-standardized incidence ratio of malignancies calculated using Cancer Statistics 2016 and 2017 by the National Cancer Registry (Ministry of Health, Labor and Welfare; Japan) was 22.4, the malignancy rate in anti-NXP2 antibody-positive adults was definitely high. In contrast, only one (3.4%) juvenile patient was diagnosed with mature B cell lymphoma at the time of the PM diagnosis.

### Relative factors of extensive muscle weakness

Thirty-six (47.4%) patients with muscle weakness extending to the distal limbs showed equal elevation of serum CK levels (median [IRs], 2205 [820–5986] U/L), while 40 patients with muscle weakness in the proximal limbs showed equal elevation of serum CK levels (3543 [1470–7247] U/L; *P* = 0.79 by Mann–Whitney *U* test). The frequency of neck muscle weakness was significantly higher in patients with muscle weakness extending to the distal limbs than patients without muscle weakness in the distal limbs (75% vs. 45%; *P* = 0.01 by Fisher’s exact test]). Among patients with DM-suggestive histopathological findings on muscle biopsy, microinfarction was more frequently observed in patients with extended muscle weakness than in the other patients (28.1% vs. 6.3%; *P* = 0.043 by Fisher’s exact test).

## Discussion

The present study revealed that 35.5% of all patients with anti-NXP2 antibody-positive IIMs present with PM without DM-specific skin manifestations and that 70.4% of these patients had nonspecific skin manifestations. Adult patients tended to present with PM phenotype more frequently than juvenile patients. These patients would have been diagnosed with PM according to the 2017 EULAR/ACR classification criteria for IIMs. However, the results of this study suggest that these patients should be treated for DM phenotype, with consideration to the possibility of complications, including internal malignancy and ILD, especially in adult patients.

Although anti-NXP2 antibody and other MSAs such as anti-TIF1γ, anti-Mi-2, anti-MDA5, and anti-SAE antibodies are reported to be strongly associated with DM-specific rashes (8, 9), a European cohort study revealed that anti-TIF1γ, anti-MDA5, and anti-SAE antibodies showed a stronger correlation with DM-specific rashes, with odds ratios (ORs) with 95% confidence intervals (CIs) of 42.68 (17.22–105.83), 43.12 (5.76–322.62) and 42.04 (10.0–175.15), respectively, than anti-NXP2 antibody (OR with 95% CI, 7.70 [3.29–17.29]) (8). Another study showed that 10%–12.5% patients with anti-NXP2 antibody were diagnosed with PM (6), while almost all the patients with anti-TIF1γ and anti-SAE antibodies were diagnosed with DM (9). In our previous study, anti-NXP2 antibody was frequently observed in cases of DMSD, which is defined as DM based on histological features of muscle biopsies, but without any skin manifestations (7). The present study also showed that 35.5% anti-NXP2 antibody-positive patients were classified as having PM according to the 2017 EULAR/ACR classification criteria for IIMs. Moreover, there were PM phenotype patients without any other skin manifestations, including periungual erythema and facial erythema, who could be diagnosed with DMSD.

It has been reported that 10%–37% and 35%–36% anti-NXP2 antibody-positive patients have complications such as subcutaneous calcinosis and edema, respectively (3, 5, 10, 11). The present study also showed that 11.6% and 26.3% patients had subcutaneous calcinosis and edema, respectively, which represented lower frequencies than those reported in previous studies. The patients in this study were all Japanese; therefore, these differences may be partially explained by the genetic background and/or accessibility of medical specialists in Japan.

The muscular manifestation of IIMs with anti-NXP2 antibody is known to be severe, with a high incidence of dysphagia (5, 8, 10) and muscle weakness of the distal extremities (5). The present study also showed that almost half of the patients had dysphagia and muscle weakness of the neck and distal extremities. We also found that the extent of muscle weakness was not related to serum CK levels. To the best of our knowledge, no previous study has reported this point. Our results from another study indicated that microinfarction, but not PFA, was a predominant muscle pathology in anti-NXP2 antibody-positive IIMs compared to other MSA-related IIMs (submitted). The present study revealed that microinfarction, detected on the muscle biopsy, was associated with the extent of their muscle weakness. In a previous study, muscle ischemic findings were often observed in patients with anti-NXP2 antibody-positive juvenile DM and these patients tended to develop more severe myopathies than others without muscle ischemia (12). These results suggest that muscle ischemia, but not muscle inflammation, may be directly correlated with muscle weakness, as shown in a murine model of spontaneous autoimmune myositis in major histocompatibility complex class I-transgenic mice due to endoplasmic reticulum stress, which could be caused by ischemia (13).

The clinical characteristics of juvenile and adult patients were not significantly different, except for the frequency of DM-specific skin manifestations. It has been reported that juvenile DM patients more frequently present with calcinosis and less frequently present with ILD and internal malignancy than adult patients (14). These trends were also observed in the present study. This study also confirmed that adult patients with anti-NXP2 antibody-positive IIMs had a higher prevalence of malignancy than the general population. The complication rate of malignancy has previously been reported to be 9.0%–37.5% (5, 6, 10). Collectively, the presence of anti-NXP2 antibody, as well as anti-TIF1γ antibody, may enhance the risk of malignancy.

In this study, the ANA titer was low (<1:80) in half of the patients with anti-NXP2 Antibody. This result suggested that the anti-NXP2 antibody titer was relatively low, which may explained that the sensitivity of a commercial blot assay kit for detecting anti-NXP2 antibody was low in a previous report (15).

The limitations of this study include a cohort of Japanese only, although we firstly demonstrated the clinical characteristics of juvenile and adult patients with anti-NXP2 Antibody-positive IIMs. Large populations lacking DM-specific skin manifestations in adult patients, and extensive muscle weakness as correlated with ischemic histological findings in muscle biopsies were especially remarkable. Future studies are needed to evaluate and establish these characteristics in large international cohorts.

## Supporting information

Supplemental list

## Data Availability

This manuscript does not include a data set that requires deposition.

## Acknowledgements

We appreciate the help of members of the different departments in collecting patients’ clinical data, as shown in the Supplemental list.

## Notes

**Funding** Supported by KAKENHI from the Japan Society for the Promotion of Science (JSPS, 18K08263 for Naoko Okiyama).

**Competing interests** MM: Grant from Chugai Pharmaceutical Co., Ltd., UCB Japan Co. Ltd., CSL Behring, Abbvie Japan Co., Ltd., Japan Blood Products Organization, Ayumi Pharmaceutical Co., Nippon Kayaku Co., Ltd, Asahikasei Pharmaceutical Co.; Consulting fees from Daiichi Sankyo Co., Ltd. and Taisho Pharmaceutical Co., Ltd.; and lecture fees from SD K.K.

### Competing Interest Statement

Masaaki Mori: Grant from Chugai Pharmaceutical Co., Ltd., UCB Japan Co. Ltd., CSL Behring, Abbvie Japan Co., Ltd., Japan Blood Products Organization, Ayumi Pharmaceutical Co., Nippon Kayaku Co., Ltd, Asahikasei Pharmaceutical Co.; Consulting fees from Daiichi Sankyo Co., Ltd. and Taisho Pharmaceutical Co., Ltd.; and lecture fees from SD K.K.

### Funding Statement

Supported by KAKENHI from the Japan Society for the Promotion of Science (JSPS, 18K08263 for Naoko Okiyama).

### Author Declarations

This study was approved by the ethics committee of University of Tsukuba Hospital (H29-111) in accordance with the Helsinki Declaration.

