## Supplemental list for "Anti-nuclear matrix protein 2 antibody-positive idiopathic inflammatory myopathies represent extensive myositis without dermatomyositis-specific rash"

Ayaka Hiraki (Hitachi General Hospital); Naoki Suzuki (Tohoku University); Fusayo Ikeda (Kinki University); Tadamichi Kasuya and Fumiko Oda, Shunjiro Kurihara, and Hitoshi Ogata (Chiba University); Ichiro Mori (Gifu University); Hiroyuki Ishida (Kyoto City Hospital); Goichi Kageyama (Hyogo Prefectural Amagasaki General Medical Center); Tomohiro Ogawa (Dokkyo Medical University); Rina Fujita and Masaru Matsui (Japanese Red Cross Otsu Hospital); Akihiro Kitamura (Shiga Medical University); Natsuki Shima (Jichi Medical University); Ryosuke Oda (Sapporo Medical University); Katsuhiko Yoneda (Shinko Memorial Hospital); Shohei Fujita (Kobe City Medical Center General Hospital); Takashi Nagai and Shinji Naito (Tokushima University); Yumiko Nakano (Kitano Hospital); Naoya Kamimura (National Hospital Organization Yokohama Medical Center); Ryota Naito (Kyoto University); Atsushi Katayama (Toyooka Hospital); Akira Yoshida, Akitsu Yoshida, and Ryo Rokutanda (Kameda General Hospital); Tsuyoshi Yoshida (Chikamori Hospital); Asami Ohara, Toaki Kohagura and Haruna Kanaseko (Aichi Children’s Health And Medical Center); Eriko Takemura (Otemae Hospital); Ai Onishi and Ren Hayasibe (Yokohama City Hospital); Kinya Matsuo (Yamaguchi University); Koichi Kitamoto (Tottori University); Satoko Yamaguchi (Tenri Hospital); Chiyu Nemoto (Matsudo City General Hospital); Yukinobu Nakagawa (Osaka University); Ryohei Komaki (Kobe University); Shoichiro Kusunoki and Nozomi Tawara (Kumamoto University); Yuko Sugita (Osama Medical University); Toshitaka Kizawa (Sapporo Hokushin Hospital); Motonori Takamiya (Kagawa Prefectural Center Hospital); Ko Matsuo (Japanese Red Cross Ise Hospital); Toyoki Nishimura (Miyazaki University); Emi Ishibazawa (Asahikawa Medical University); Yuki Fujii (Teikyo University); Mariko Nikaido (Yamagata University); Nobuyuki Eura (Nara Prefectural Medical University); Mutsuki Takeda (Yokohama Sakae Kyosai Hospital), Yumiko Sugiyama (Yokohama Minami Kyosai Hospital); Hiro Tani (Hiroshima University); Hiroyuki Koizumi (Teikyo University School of medicine University Hospital, Mizonokuchi); Shinichiro Omura (Seirei Hamamatsu General Hospital)
